# From acute injury to chronic comorbidity: Interrupted time series modeling of traumatic brain injury impact among post-9/11 veterans

**DOI:** 10.1101/2025.05.16.25327801

**Authors:** Mustafa Ozmen, Shashank Vadlamani, James J Gugger, Megan Amuan, Amanda Cheney, Ramon Diaz-Arrastia, Mary Jo Pugh, Eamonn Kennedy

## Abstract

Traumatic brain injury (TBI) is associated with a variety of adverse health outcomes that display complex behavior over time. The objective of this study was to investigate both the early and late health impacts of TBI within a single framework. This study evaluated TBI associations among a cohort of post-9/11 Veterans with TBI documented between 2008 and 2017 in Veteran Health Administration (VHA) records. The cohort included 108,408 post-9/11 Veterans with any history of TBI documentation between 2008-2017 who were demographically matched with 108,408 TBI negative controls. Interrupted time series (ITS) models were used to fit the prevalence of comorbidities over time (±6 years from index date, i.e. date of first TBI). Three ITS measures were modeled for each comorbidity: 1) The incidence rate (IR) in the month of TBI index date, 2) The incidence rate ratio (IRR) between TBI and control groups in the month of index date, and 3) Long-term changes in year-over-year diagnosis rates, i.e. the annual incidence rate difference (IRD) before vs. after index date. Overall, TBI was associated with conditions related to somatic, cognitive, and psychological outcomes including headache, cognitive dysfunction, and PTSD. Neurological events were found to be elevated within the month of TBI documentation. Conditions with the largest IR were post-traumatic stress disorder (PTSD) (+29%, p<0.001), headache (+22%, p<0.001), and adjustment disorder (+22%, p<0.001). Conditions with the highest IRR across TBI and control groups were cognitive dysfunction (474, p<0.001), vestibular dysfunction (137, p<0.001), and stroke (72, p<0.001). Long term, the conditions with the highest IRD were substance use disorders (p<0.001) and mental health conditions (p<0.001). This work demonstrates how ITS modeling can help bridge traditional divides between early and late paradigms of TBI investigation to help inform research and care for Veterans living with TBI.

## Introduction

TBI is a prevalent and heterogeneous condition that is associated with a variety of adverse outcomes across mental and physical health domains [1]. TBI has been traditionally classified as mild, moderate, or severe [2] based on clinical features at the time of injury, such as the presence and duration of unconsciousness, post-traumatic amnesia, and Glasgow Coma Scale (GCS) scores [3]. Recent evidence supports that distinct clinical presentations or phenotypes exist within the TBI population that depend on injury-related factors that are not well captured by existing severity classifications [4]. Cumulatively, this heterogeneity creates challenges and opportunities for the optimization of care and recovery after injury [5]. While good recovery is expected for most cases following TBI, a variety of adverse consequences of TBI can emerge and persist long after injury [4]. The emergence of TBI-related concerns over time is complex, and distinct phenotypes can emerge and diverge, leading to challenges including polypharmacy among survivors of TBI [6]. Therefore, it is crucial to use methods that can capture multiple disease trajectories and diagnoses following brain injury [7].

Identifying TBI comorbidity is a core concern for military health because Service Members and Veterans (SMV) experience elevated rates of TBI and large numbers of TBI-related health concerns. More than 505,000 SMV who served in Iraq and Afghanistan have been diagnosed with TBI [8], and history of TBI has been linked to a range of military health concerns including post-traumatic stress disorder (PTSD), chronic pain, headache [9], sleep problems, epilepsy, stroke, and cardiovascular disease [10,11].

TBI has traditionally been considered an acute event with a finite period but is increasingly recognized as a chronic condition with wide-ranging effects [12,13]. Therefore, both the acute and chronic impacts of TBI should be considered together to better understand TBI health outcomes. However, to date, studies have typically only considered the basic properties of comorbidity risk following injury such as prevalence to evaluate TBI health impacts, but many other metrics can be obtained, each with unique advantages and clinical implications. For example, modeling the immediate increase in diagnoses within just the month of injury documentation could help prioritize acute risks, while difference measures comparing diagnosis in the years preceding/following TBI could identify latent, long-term health changes.

The purpose of this study was to leverage interrupted time series (ITS) modeling of TBI comorbidities among Post-9/11 Veterans to provide a comprehensive picture of TBI comorbidity over time [14]. ITS is a useful approach for understanding how events like TBI can disrupt, redirect, or accelerate the development of health concerns. ITS metrics were selected to yield insights into the timing, intensity, and persistence of health status changes related to TBI. Pre-and post-exposure trends of each comorbidity were compared across TBI and control groups, interrupted at the time of primary exposure, i.e. the index date.

This study focused on three measures: **1)** Monthly incidence rate (IR) in the month of injury. **2)** Early effects of TBI measured by the monthly incidence rate ratio (IRR) between TBI and Control Group in the month of injury. **3)** Chronic long-term trend changes in diagnoses of comorbidities measured by the annual incidence rate difference (IRD) before-to-after injury. A conceptual diagram illustrating these three measures is provided in Fig 1. We hypothesized that compared to matched controls, Veterans with history of TBI would show distinct health trajectories after the date of injury (index date) across IR, IRR, and IRD metrics up to 6 years after first documented TBI.

**Fig 1:**
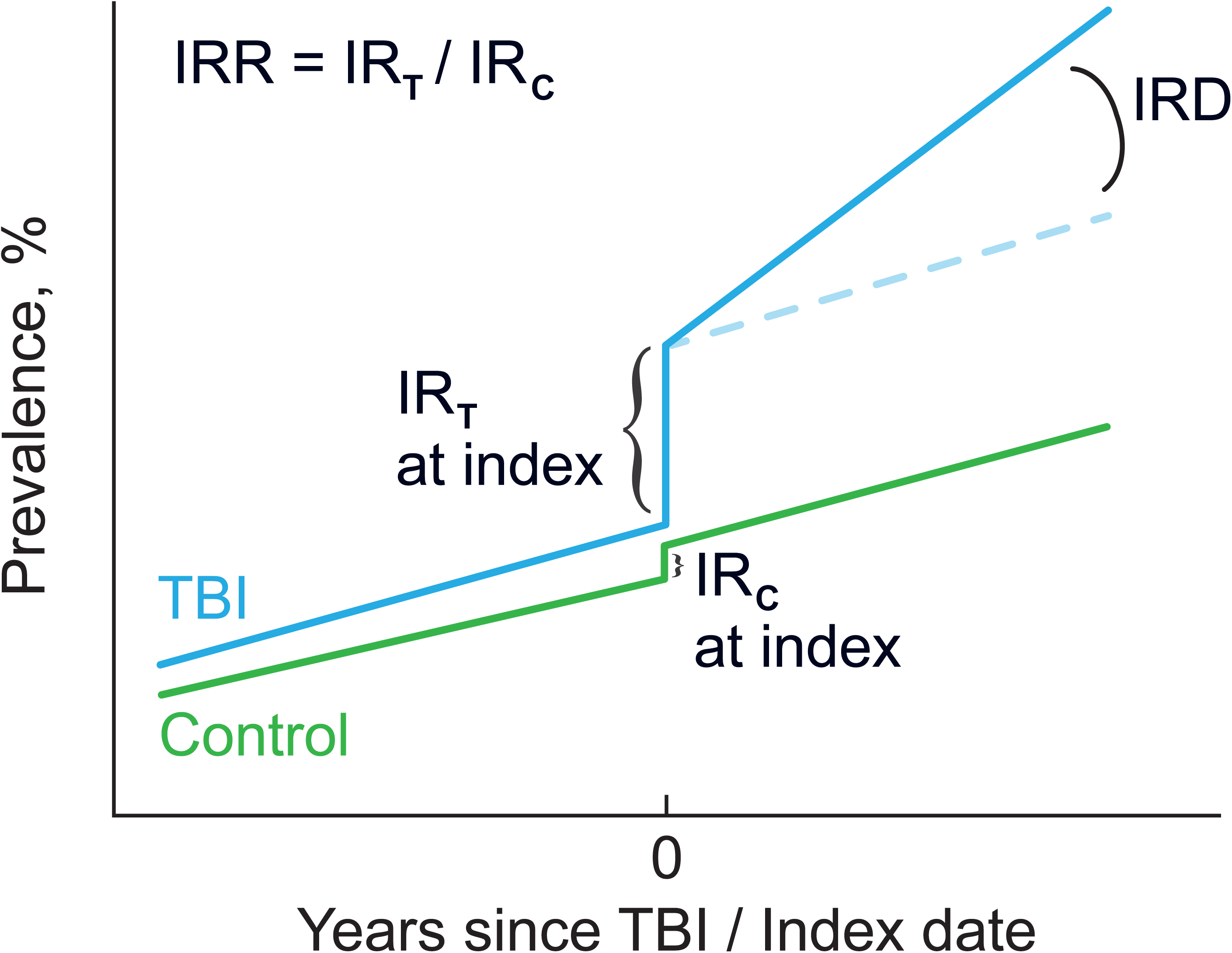
Conceptual diagram illustrating ITS measures. (IR: Monthly incidence rate at TBI index, IRR: Monthly incidence rate ratio between TBI and Control Group at TBI index, IRD: Annual incidence rate difference before-to-after injury.)

## Materials and methods

### Cohort description

The original cohort included N = 910,951 Post-9/11 Veterans who entered Veteran Health Administration (VHA) care between October 1, 2001, and September 30, 2014. To exclude individuals who minimally used services, we only included Veterans who engaged in 2 or more years of VHA care (inpatient, outpatient, or pharmacy) before September 30, 2014. Separately, to allow for a 6-year observation time window both before and after TBI diagnosis date (index date) and to account for implementation of the VA TBI screening and comprehensive evaluation process in 2007, only Veterans with a first TBI diagnosis between the years 2008 and 2017 were included. These two criteria ensured that there was adequate information per individual, as well as adequate observation time per individual across a 6-year time window. For this cohort, we obtained data from the Defense Health Agency (DHA) and VHA (Veterans Health Administration) through DaVINCI (Veterans Affairs Informatics and Computing Infrastructure) in addition to community care data (FY99-FY23) to examine health status over time. The exposed cohort consisted of 108,408 Veterans with TBI of any severity. All TBI cases were exactly matched 1:1 with unique controls on age, sex, race, and ethnicity, drawing from TBI negative cases in the original cohort. The index date for each Veteran in controls was set equal to the index date of their matched Veteran with TBI.

### TBI classification

To classify the primary exposure of TBI, we used established severity definitions [15]: None, mild, moderate/severe, and penetrating injury. Evidence of TBI was drawn from multiple sources, including the DoD Trauma Registry, VHA inpatient and outpatient data, and self-report on the VA Comprehensive TBI Evaluation (CTBIE). TBI severity was based on ICD-9/10 codes and the CTBIE clinical interview [16]. If a veteran met criteria for more than one TBI severity, the higher severity level was assigned. Classification of No TBI required no evidence of any classified or unclassified TBI. In the CTBIE, a loss of consciousness (LOC) of 0-30 minutes, or an alteration of consciousness or post-traumatic amnesia (AOC/PTA) = 0-1 day is classified as mild TBI. An LOC > 30 min, AOC/PTA > 1 day is classified as moderate/severe. Penetrating TBI classified injuries where objects entered the brain. Using these definitions, groups were classified according to lifetime TBI status: 1) No Lifetime TBI, 2) At least one lifetime TBI, which included unclassified, mild, moderate/severe and penetrating TBI severity. Case severity was then further stratified as either mild TBI (the majority of cases) or Moderate or above severity, excluding unclassified TBI.

### TBI comorbidities

Selection of comorbidities was based on the frequency of conditions within the Veteran community and priority of their relationship to TBI based on expert consensus [17]. We defined 65 diagnostic variables according to the presence of related ICD-9/10 codes in DoD/VHA data. Veterans were only considered to have a diagnosis if their health record contained two or more relevant codes separated by one ITS time step (30 days) [18]. The variables included objective measures, disease diagnosis, and diagnosis of symptoms. Conditions that had no evidence of diagnosis in either group before index date were excluded from ITS analysis.

### Demographics and military measures

Demographic data and military characteristics were obtained from VHA administrative data. Military descriptive variables included active-duty status, rank, and deployment history including time since last deployment. Individuals with more than one date of deployment were identified as having multiple deployments.

### Statistical analysis

All analysis were conducted in Python 3.11. Descriptive statistics were used to summarize demographic, clinical, and psychosocial characteristics. ITS models were used to assess temporal trends in diagnosis with respect to index date. ITS models, metrics, and statistical tests were conducted according to guidelines (see Section: Criteria, below).

### Data availability and access statement

Raw data is not available because it is identifying. Some deidentified data is available upon reasonable request pending appropriate study approvals and data transfer agreements between participating institutions. Code used for analysis is available upon request.

### Standard protocol approvals, registrations, and patient consents

The overall study was approved by the institutional review board (IRB) of the University of Utah with a waiver of informed consent. This study was limited to secondary deidentified data analysis and was designated as non-human subject’s research. Following ethics approval obtained for this study on June 9^th^ 2023, data were accessed for this study on August 22^nd^, 2023.

### Interrupted Time Series

ITS is a method to analyze data collected over time when an event, exposure, or intervention disrupts or creates a change in behavior at a specific point in time [14]. The analysis requires that data is available both before and after the event (first TBI date) per person, so that the impact of the interrupt event on the underlying secular data can be determined (Fig 1). The ITS model used the equation:

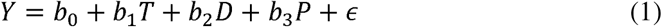

In the equation, Y is outcome variable which is the prevalence for a given comorbidity. T is a continuous time variable (-6 years to +6 years centered at TBI index date). D is the interrupt variable (i.e., a variable indicating TBI has occurred), D=0 before index date, D=1 after index date. P is the time passed since intervention (P=0 before index date, P=T after index date, and c is zero centered Gaussian random error.

#### Metrics

The first metric modeled by ITS was the pre-injury prevalence of conditions. Prevalence was calculated as the percentage of each group who ever met diagnostic criteria for the condition up until the month of TBI injury / index date. Incidence rate (IR) was calculated as the number of new cases of a disease or condition that occurred during the month of first injury / index date, expressed as a percent per month. The incidence rate ratio (IRR) was calculated as the ratio of incidence rates between TBI and Control groups in the month of first TBI, i.e. IR_t_/IR_c_. The final metric was the annual incidence rate difference (IRD) which was calculated as the percentage difference in incidence rates before to after index date for each group.

#### Criteria

ITS models are useful in health data assessment, but a systematic approach and careful consideration of ITS criteria is required. Ramsay et. al. proposed quality criteria for ITS design [19]. The first criterion concerns the independence of the exposure. The current study meets this criterion because most of the comorbidities selected have minimal influence on the risk for subsequent TBI except for substance use disorders which is discussed as a limitation (see Limitations). The second criterion is whether the occurrence of the exposure/event is likely to impact data collection. At the time of TBI documentation, additional visits to health professionals and clinics might occur. Therefore, there is some risk for influence of data collection on results, which is a noted limitation (see Limitations). The third and fourth criteria relate to the unbiased and reliable measurement of the primary outcomes. For this we can refer to the good positive predictive values of TBI and other measurements from previous studies conducted using the same cohort (see Methods: TBI classification). The fifth criterion concerns whether there is adequate coverage of participants at each point across time. The original data is collected between 1999 and 2023, so including all subjects with index dates across this range would be inadequate for follow up. Instead, to meet this criterion, only participants with the first TBI documented between the years 2008-2017 were included to ensure coverage across a 6-year time window.

## Results

After exclusion criteria, the total sample was n = 216,816, including 108,408 Veterans with lifetime history of TBI of any severity demographically matched 1:1 with controls. Summary statistics for demographic, military, and injury measures are shown in Table 1. The TBI and matched control groups showed similar demographic and military characteristics. The mean age at time of TBI documentation (index date) was 30 years old and 7.3% of subjects were female. More than half of the total sample had multiple prior deployments. In the TBI group, 67.5% had mild TBI, 14.0% had moderate/severe, 2.6% had penetrating TBI, and 15.9% had unclassified TBI severity.

**Table 1:**
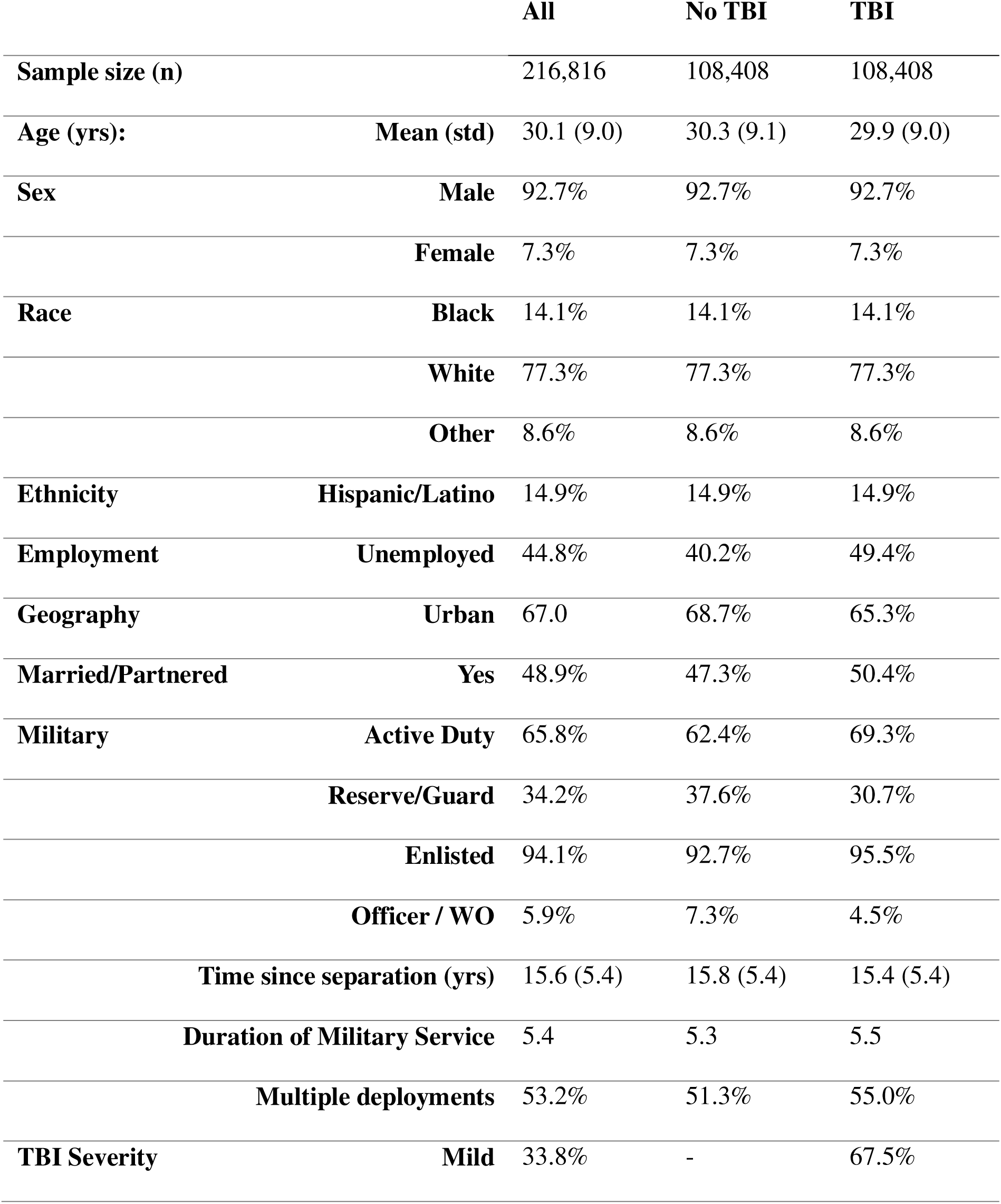
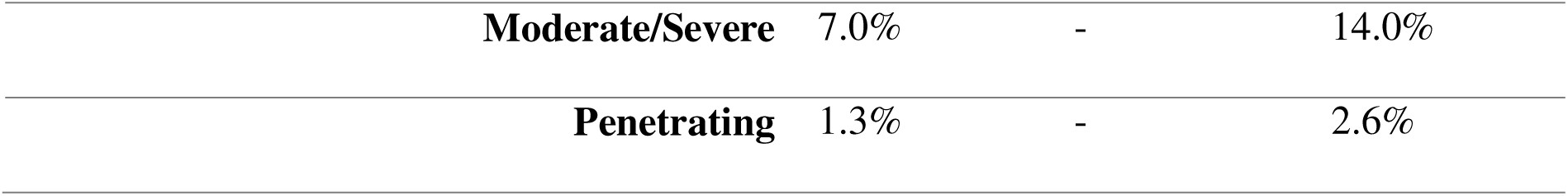
Characteristics of the cohort.

|  |  | **All** | **No TBI** | **TBI** |
| --- | --- | --- | --- | --- |
| **Sample size (n)** |  | 216,816 | 108,408 | 108,408 |
| **Age (yrs):** | **Mean (std)** | 30.1 (9.0) | 30.3 (9.1) | 29.9 (9.0) |
| **Sex** | **Male** | 92.7% | 92.7% | 92.7% |
|  | **Female** | 7.3% | 7.3% | 7.3% |
| **Race** | **Black** | 14.1% | 14.1% | 14.1% |
|  | **White** | 77.3% | 77.3% | 77.3% |
|  | **Other** | 8.6% | 8.6% | 8.6% |
| **Ethnicity** | **Hispanic/Latino** | 14.9% | 14.9% | 14.9% |
| **Employment** | **Unemployed** | 44.8% | 40.2% | 49.4% |
| **Geography** | **Urban** | 67.0 | 68.7% | 65.3% |
| **Married/Partnered** | **Yes** | 48.9% | 47.3% | 50.4% |
| **Military** | **Active Duty** | 65.8% | 62.4% | 69.3% |
|  | **Reserve/Guard** | 34.2% | 37.6% | 30.7% |
|  | **Enlisted** | 94.1% | 92.7% | 95.5% |
|  | **Officer / WO** | 5.9% | 7.3% | 4.5% |
|  | **Time since separation (yrs)** | 15.6 (5.4) | 15.8 (5.4) | 15.4 (5.4) |
|  | **Duration of Military Service** | 5.4 | 5.3 | 5.5 |
|  | **Multiple deployments** | 53.2% | 51.3% | 55.0% |
| **TBI Severity** | **Mild** | 33.8% | - | 67.5% |

|  |  |  |  |
| --- | --- | --- | --- |
| **Moderate/Severe** | 7.0% | - | 14.0% |
| **Penetrating** | 1.3% | - | 2.6% |

Fig 2 shows the prevalence of four comorbidities over time, with the index date set to zero years. The TBI and control groups also showed comparable incidence rates for most comorbidities up to the index date. Fig 2a shows the prevalence of attention disorders over time, which have been previously associated with TBI [20,21]. There is a clear temporal relationship between Attention disorder diagnosis and the date of the first TBI. Before the index date, prevalence is low, and TBI severity groups show similar diagnosis rates. At the time of the index date, there is a large increase in new cases of attention disorders for TBI groups but not among controls (green).

**Fig 2:**
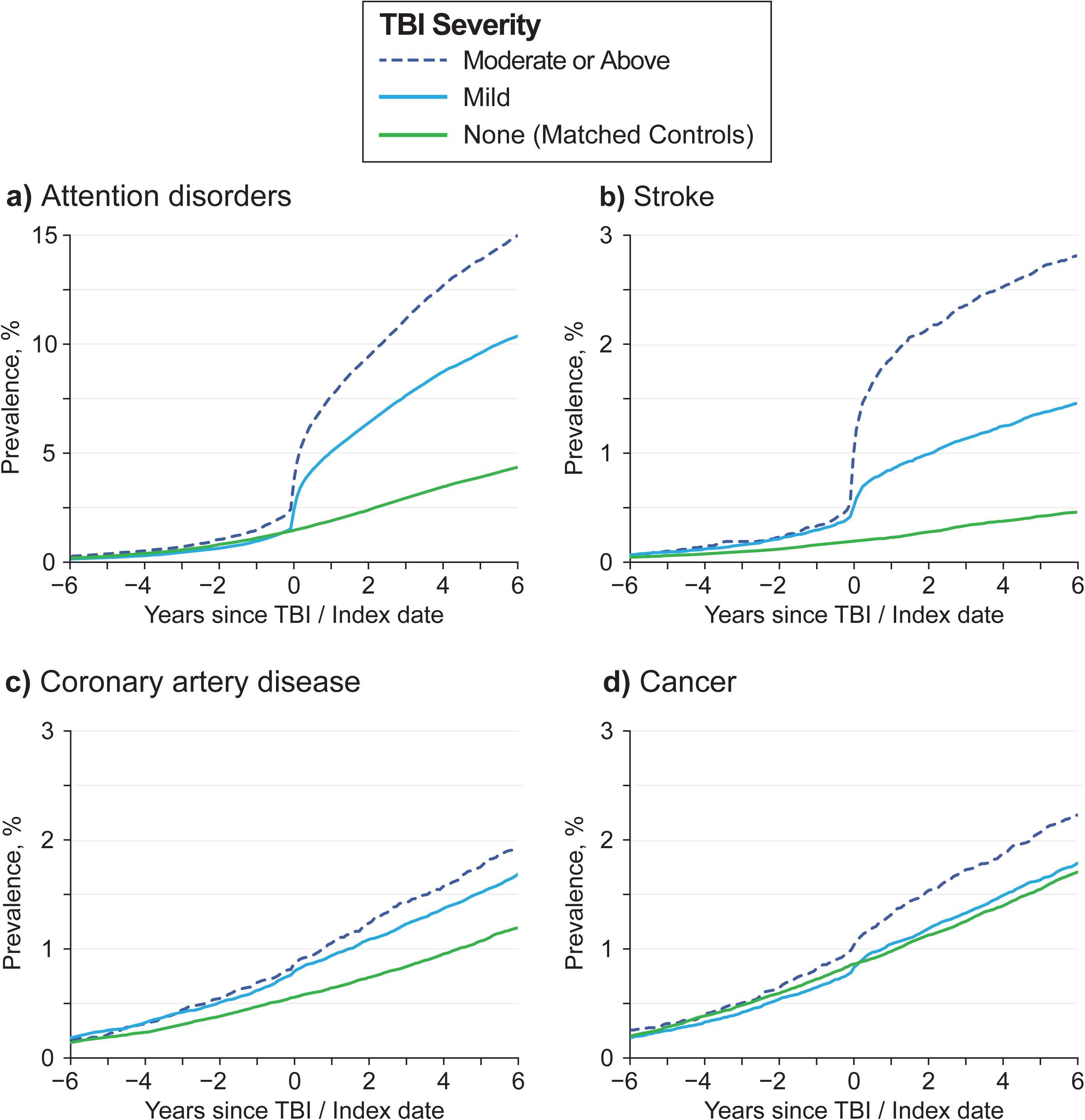
Prevalence of Comorbidities. Plots of the prevalences for four conditions +/- 6 years from date of first TBI, shown for three groups: matched control (green), mild TBI (cyan) and moderate or above severity TBI (dotted blue). Attention disorders **(a)** and stroke **(b)** show clear increases in prevalence both at and after the index date among the TBI groups, but not among controls. Conversely, coronary artery disease **(c)** and cancer diagnosis **(d)** do not show a clear trend at or after the first index date, indicating minimal temporal association with TBI.

Fig 2b shows a similar pattern with the prevalence of stroke with respect to index date. The higher incidence rate (slope) for the TBI group compared to controls after TBI index date indicates the chronic effects of TBI are associated with a long-term risk for stroke for many years after the first TBI documentation. Together, these data suggest both acute and chronic relationships between TBI and these conditions. Fig 2c-d shows the prevalence of coronary artery disease (CAD) and cancer over time. There is no clear temporal relationship between CAD or cancer diagnosis and the date of first TBI. Together, these data suggest no clear acute and chronic relationships between TBI and CAD or cancer diagnosis.

Fig 3 shows the ITS model fits of the prevalence of sixteen conditions over time for each group. Relative to the TBI index date, a long-term trend of elevated diagnosis rates among the TBI group is evident over many years for multiple conditions. Conditions such as PTSD and sleep disorders show a large change in diagnosis rates coinciding with the time of first TBI diagnosis. Other conditions such as cannabis abuse do not show a clear change around the time of TBI diagnosis, but instead display chronic increases in the years following TBI. Some conditions such as bipolar disorder showed both a large change at the index date as well as a chronic long-term increase in diagnosis following the index date.

**Fig 3:**
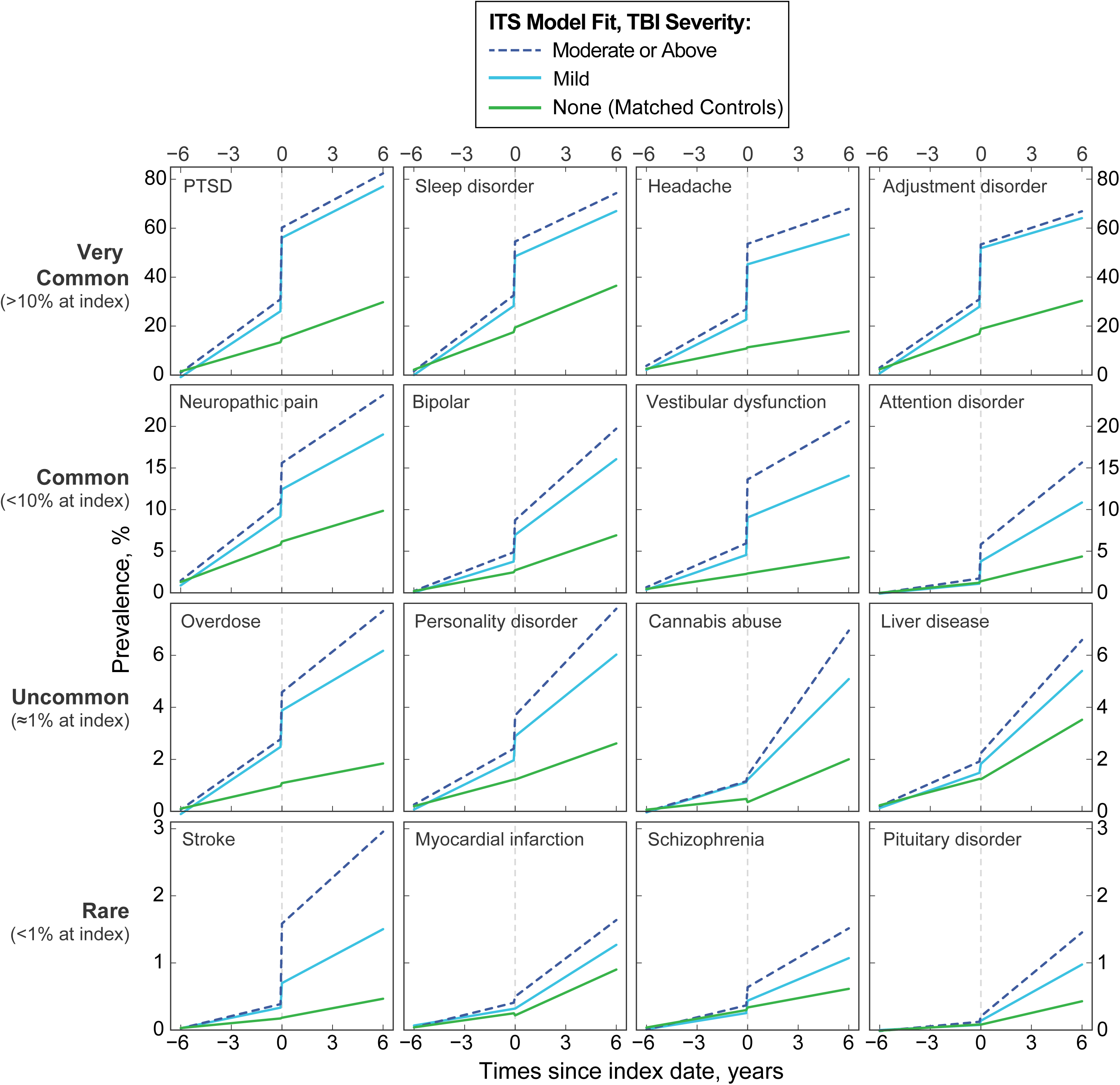
ITS model fits of the prevalence for sixteen conditions. Prevalences are shown over time matched for three groups: control (green), mild TBI (cyan) and moderate or above severity TBI (dotted blue). Comorbidities are shown in descending levels of prevalence. Compared to the TBI group trends, similar patterns were typically not present among controls.

To provide one example of ITS metrics, the results of the ITS fit of attention disorders are shown in Table 2. Key parameters are listed alongside descriptions and values for attention disorders for TBI groups. The impact of TBI severity is evident by comparing values across groups. The IRR in the month of first TBI with respect to the same time period among controls indicates the immediate, relative impact of TBI. Comparing IRRs of mild TBI and moderate or above severity TBI groups also indicated stronger association for higher severity injury. Annual IRD for mild and moderate or higher severity TBI groups was higher than the controls.

**Table 2:** Descriptions and measures of key ITS features with values for attention disorder.

| Property | Description | Values for Attention Disorders |  |
| --- | --- | --- | --- |
|  |  | Mild TBI | Moderate or above TBI |
| **Prevalence** | Percentage ever diagnosed before first TBI in TBI group. | 1.86% | 2.80% |
| **IR** | Percentage newly diagnosed in the month of first TBI in TBI group. | 3.87% per month | 5.30% per month |
| **IRR** | Ratio of percentage ever diagnosed across TBI and Control Groups in the month of first TBI. | 24.8 | 34.0 |
| **IR after index for TBI group** | Annual rate of new diagnosis among the TBI group after index date | 1.62% per year | 2.23% per year |
| **IR after index for controls** | Annual rate of new diagnosis among the Control group after index date | 0.63% per year | 0.63% per year |
| **IRD for TBI group** | Annual Incidence Rate change<br>before/after index date in TBI<br>group. | 1.38% per year | 1.88% per year |
| **IRD for control**<br>**group** | Annual Incidence Rate change<br>before/after index date in Control<br>group. | 0.37% per year | 0.37% per year |

Comprehensive metrics for all conditions are available in S1 Table.

Extending beyond single examples, Table 3 shows ITS metrics for 27 comorbidities fit for all Veterans with history of TBI. These comorbidities were selected according to the union of the Top 10 variables in the TBI group for Prevalence, IR, IRR, and IRD. Pain, PTSD and sleep disorders were among the most prevalent conditions. PTSD, headache, adjustment disorder, depression, and sleep disorders had the highest IR within the month of index date. IRRs were highest for neurological concerns, including cognitive dysfunction, vestibular dysfunction and stroke, and headache.

**Table 3:** TBI group ITS features for 27 comorbidities, shown ranked by percentage of Veterans with the diagnosis prior to month of injury (prevalence).

**ALL TBI Group ITS Model Estimates**
|  | **Prevalence** | **IR** | **IRR** | **IRD** |
| --- | --- | --- | --- | --- |
| **Condition** | *Pre-injury, %* | *TBI, % / month* | *TBI:Control* | *before/after TBI, % / year* |
| Other Pain | **56.2** | 9.4 | 7 | -5.3 |
| Back Pain | **49.9** | 13.0 | 9 | -4.2 |
| PTSD | **35.0** | **29.0** | 23 | -1.4 |
| Sleep Disorder | **35.0** | **19.7** | 12 | -1.9 |
| Smoker | **34.4** | 7.8 | 6 | -3.3 |
| Adjustment Disorder | 34.1 | **22.4** | 12 | -2.7 |
| Depression | 30.8 | **18.4** | 15 | -0.80 |
| Headache | 27.5 | **22.4** | **51** | -1.5 |
| Anxiety | 26.0 | 15.2 | 12 | -0.67 |
| Hypercholesterolemia | 18.7 | 5.5 | 6 | 0.37 |
| Neck Pain | 18 | 7.7 | 23 | -0.61 |
| OSA | 10.4 | 4.1 | 13 | **1.0** |
| Vestibular Dysfunction | 5.7 | 4.8 | **137** | 0.13 |
| Bipolar | 5.4 | 3.3 | 18 | **0.84** |
| Cognitive Dysfunction | 4.4 | 10.1 | **474** | **0.58** |
| Personality Disorder | 2.6 | 0.97 | * | 0.2 |
| Opioid Abuse | 1.9 | 1.1 | 14 | 0.31 |
| Any attention disorder | 1.9 | 2.9 | 22 | **0.98** |
| Liver | 1.8 | 0.31 | * | 0.38 |
| Cannabis Abuse | 1.5 | 0.15 | * | **0.52** |
| Dysphagia | 1.3 | 0.55 | 23 | 0.14 |
| Psychosis | 1.2 | 0.78 | 18 | 0.28 |
| Stroke | 0.48 | 0.49 | **72** | 0.08 |
| Conversion Disorder | 0.10 | 0.07 | **31** | 0.02 |
| MS | 0.10 | 0.04 | 30 | 0.00 |
| Encephalopathy | 0.04 | 0.07 | 27 | 0.02 |
| Anoxic Brain | 0.02 | 0.04 | 16 | 0.00 |

The difference in incidence rate before and after TBI (IRD) was highest for OSA, attention disorder, bipolar disorder, cognitive dysfunction, and cannabis abuse. Other conditions with high IRD included mental health and substance abuse related concerns such as opioid abuse and psychosis. The full range of ITS measures estimated for both the TBI and control groups are available in S1 Table.

The top 5 conditions with the largest values are highlighted in bold. “*” indicates ITS model fit error. (IR: Incidence Rate; IRR: Incidence Rate Ratio; IRD: Incidence Rate Difference; OSA: Obstructed Sleep Apnea; MS: Multiple Sclerosis)

The top 10 comorbidities for each metric are visualized in Fig 4 (a: Prevalence, b: IR, c: IRD, d: IRR). In Fig 4a, pain was highly prevalent among both TBI severity groups (back pain, headache, neck pain, other pain,) alongside PTSD, headache, anxiety, depression, and sleep issues. These conditions closely mirror the polytrauma clinical triad identified in previous studies [22]. In Fig 4b, incidence rates of new conditions in the month of injury were highest for PTSD, headache, sleep problems, pain, adjustment disorder, depression, and anxiety. Fig 4c showed long term chronic increases were elevated for physical and mental health concerns as well as substance use issues including opioid and cannabis abuse. In Fig 4d, the highest IRRs were specific to neurological conditions such as cognitive and vestibular dysfunction, and stroke.

**Fig 4:**
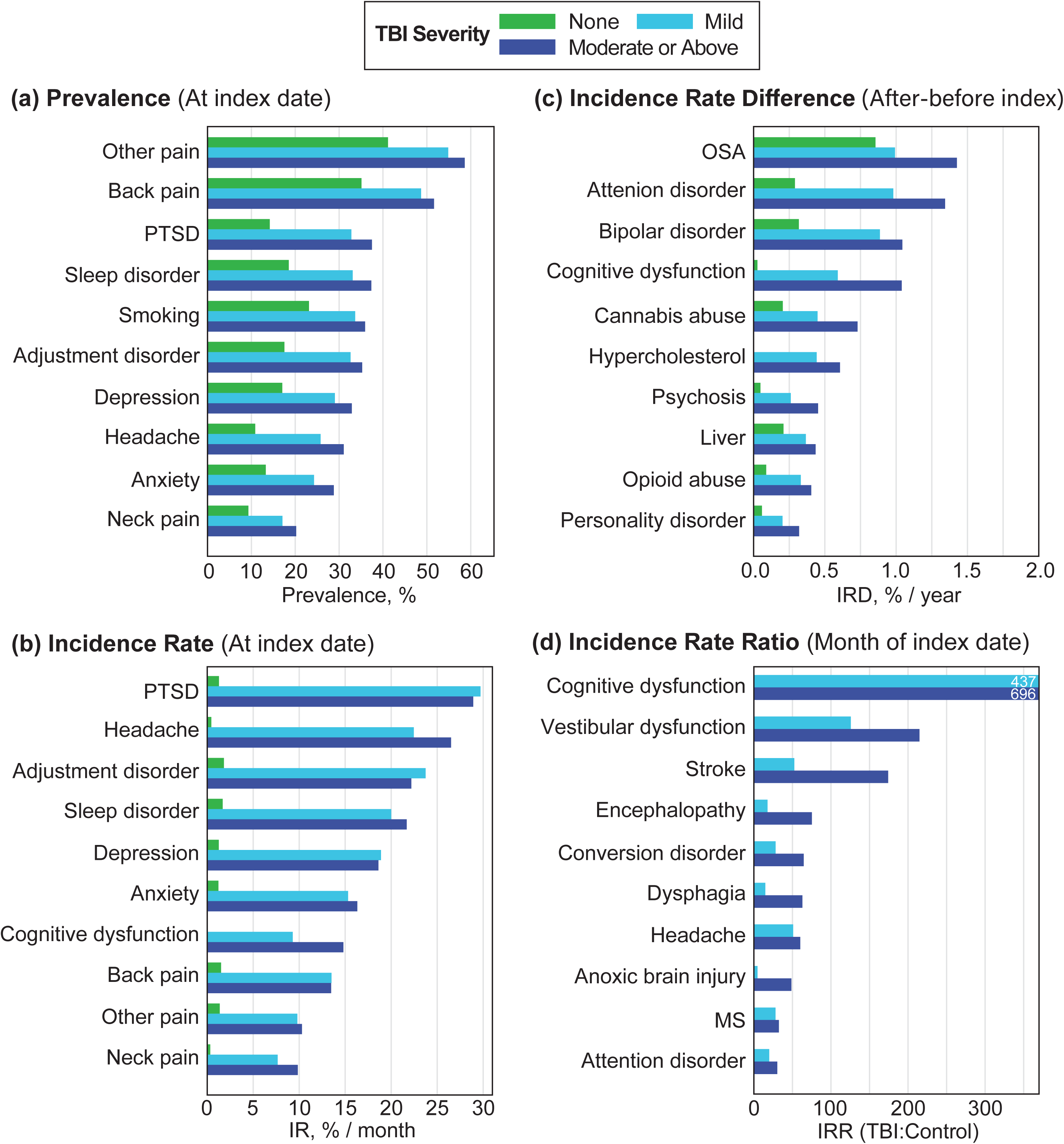
Bar graphs of the top 10 comorbidities for each measure. Conditions are ranked by a: Prevalence at TBI index date, b: Incidence rate, c: Annual Incidence Rate differences before and after TBI index date and d: Incidence Rate Ratios between TBI and Control groups.

## Discussion

TBI is associated with a variety of acute and chronic health outcomes that display complex relationships over time. To assess a broad spectrum of health risks accompanying TBI, this study investigated the early and late health impacts of TBI within a single ITS model framework [1]. ITS is a useful approach for understanding how events like TBI can disrupt, redirect, or accelerate the development of health concerns over time. Overall, TBI was strongly associated with conditions related to somatic, cognitive, and psychological outcomes, including headache, cognitive dysfunction, and PTSD [23,24]. Comparing TBI and control groups, PTSD [11], headache [9], adjustment disorder, depression [25], and sleep problems [24] were all associated with TBI. Other conditions such as cardiac diagnoses showed significant association with TBI as identified in prior work [26] but were not among the highest ranking conditions. Highest ranking conditions tended to relate to the polytrauma clinical triad, which has been previously identified in TBI phenotyping studies [4,22] with similar trends not evident among controls.

IRR assessed the degree of new TBI-specific health changes not observed in the control group. Vestibular and cognitive dysfunction showed strong IRR in the month of injury [27], which suggests an early phenotype of neurological concern. Events such as stroke and anoxic brain injury can be related complications of injury events, and these were also found to be coincident with TBI [28,29]. The relative diagnostic increase in neurological conditions following TBI aligns with the perspective of TBI as a disruptive event that can reveal latent vulnerabilities in the brain [30]. For example, preexisting subtle deficits in vestibular and cognitive dysfunction may only become clinically apparent at the time of TBI documentation [31]. The proximity of neurological diagnoses to the time of TBI documentation further suggests potential shared mechanisms such as stress and inflammatory responses [32]. Collectively, these findings suggest TBI may amplify susceptibility to a spectrum of early neurological changes.

Long term, conditions with the highest year-over-year annual increase in percentage diagnosis years after injury were predominantly mental health and substance use related comorbidities, including bipolar disorder, attention disorders, cannabis use disorder, and opioid use disorder [33]. However, the chronic rate increase of many of these conditions for the control group was also moderately elevated, suggesting underlying age-related change in risks. Our results suggest that the risk for substance misuse after TBI intensifies over time alongside cognitive, mental health, and psychiatric comorbidities that can persist and accelerate many years after injury. Mechanistically, TBI can disrupt dopaminergic and limbic pathways, which are important systems for reward processing and stress response, thereby predisposing individuals to impulsive and addictive behaviors, particularly in the presence of chronic pain [34] and stress which are common consequences of TBI [35]. These findings underscore the need for longitudinal research and treatment strategies that address a broad spectrum of risks accompanying TBI.

This work demonstrates how consideration of different time scales in a single analysis can offer opportunities to bridge acute and chronic paradigms of TBI investigation. Moreover, several temporally distinct phenotypes of complex comorbidity were identified that may benefit from clinical follow up at times of high risk for the emergence of specific TBI comorbidities.

## Limitations

Selected comorbidities showed minimal influence on the risk for subsequent TBI, with the exception of substance use disorders, which demonstrated a bidirectional relationship with TBI. It is possible that measurement bias may underly some of the TBI and control group differences. This is because TBI is a medical issue and patients with TBI may make additional visits to health professionals and clinics after their injury, increasing the probability of other diagnoses. As in all studies of electronic health records, our results are subject to measurement and documentation biases. TBI diagnosis between 2008 and 2017 were included, but the index date of TBI may not always be known or documented with certainty. The diagnosis of TBI can also reflect the date that an interview after injury was completed. There are numerous potential time series models, such as ARIMA, which are equally appropriate for analysis of these data [36]. We elected to use ITS modeling as a linear approximation because the coefficients are intuitive, and this work was primarily focused on changes at the time of TBI.

## Conclusion

TBI remains a critical risk factor for neurological disease that requires appropriate temporal modeling [37]. Using an ITS design, in this study we investigated clinically relevant temporal measures that provided several unique perspectives on TBI-comorbidity relationships. Consistent with prior work, TBI was associated with conditions related to somatic, cognitive, and psychological outcomes including headache, cognitive dysfunction, and PTSD. Neurological events were found to be elevated within the month of TBI documentation for all TBI severity groups compared to controls. A long-term pattern of elevated diagnosis was observed for mental health and substance abuse-related concerns that persisted for at least six years following injury in the TBI group.

## Transparency, rigor, and reproducibility

All codes and scripts used for this investigation are available from the corresponding author upon reasonable request.

## Conflict of interest

The authors declare no conflict of interest.

## Acknowledgments

The authors declare no acknowledgments.

## Supporting Information

**S1 Table: Detailed TBI and Control group ITS features for all (65) comorbidities.**

* = suspected model error

## Notes

### Competing Interest Statement

The authors have declared no competing interest.

### Funding Statement

This project was funded by the Congressionally Directed Medical Research Programs Epilepsy Research Program in 2023 under award EP220089.

### Author Declarations

The institutional review board (IRB) of the University of Utah waived ethical approval of this work as IRB #00163364. This study was limited to secondary deidentified data analysis and was designated as non-human subjects research. Following ethics approval obtained for this study on June 9th 2023, data were accessed for this study on August 22nd, 2023.

